# Comparison of adhesion prevention capabilities of the modified starch powder-based medical devices 4DryField^®^ PH, HaemoCer™ PLUS and StarSil^®^ in the Optimized Peritoneal Adhesion Model

**DOI:** 10.1101/2023.07.04.23292224

**Authors:** Daniel Poehnert, Lavinia Neubert, Markus Winny

## Abstract

**Background and Objectives:** The rat Optimized Peritoneal Adhesion Model (OPAM) was developed to provoke adhesion formation with high reproducibility in incidence and extent. In a recent study, the starch-based hemostats 4DryField PH and Arista AH were tested for their capabilities to prevent adhesion formation, the former one certified for adhesion prevention and hemostasis, the latter one only certified for hemostasis. As two further starch-based hemostats, i.e., HaemoCer PLUS and StarSil, have officially been certified for adhesion prevention in the meantime, the present study was conducted to examine their efficacy.

**Materials and Methods:** For this purpose, all three products were applied as a powder that was mixed *in situ* with saline solution to form a barrier gel. Adhesions were scored using the established macroscopically scoring systems by Lauder and Hoffmann, as well as histopathologically using the score by Zühlke. Animals receiving saline solution solely served as controls.

**Results:** As previously published, 4DryField PH reduced peritoneal adhesions significantly. In contrast, HaemoCer PLUS and StarSil did not lead to a statistically significant reduction of adhesion formation. When comparing 4DryField PH, HaemoCer PLUS and StarSil, 4DryField PH was significantly more effective in preventing peritoneal adhesions. The results of the macroscopic investigation were confirmed by histopathological evaluations.

**Conclusions:** Only 4DryField PH but neither HaemoCer PLUS nor StarSil were capable to effectively prevent adhesion formation, corroborating the assumption that starch-based hemostats do not generally have the capability to act as effective adhesion prevention devices.

## Introduction

Intraabdominal adhesions are a substantial inborn problem of surgery, developing after 55 –100% of all abdominal surgeries [1]. Adhesions are fibrin bands that form between damaged peritoneal surfaces normally not connected to each other, triggered on a biochemical and cellular level to repair the peritoneum, which has been damaged due to surgical trauma [2, 3]. Correspondingly, adhesions are the main cause of chronic abdominal pain, secondary female infertility, and small bowel obstructions [3-5], the latter induced by adhesions in up to 85.6% of patients [6] and with a mortality rate of up to 15% during hospitalization [7]. Furthermore, adhesions lead to a high incidence of re-admissions and re-interventions [5, 8], exacerbated by reduced visibility and difficult accessibility, increasing the risk of inadvertent enterotomies and significantly prolonging operation time [9].

Considering the severe burden imposed to patients, surgeons, hospitals and the healthcare system, manifold attempts have been made to design effective anti-adhesive agents [10]. Many of these devices act as physical barriers to separate traumatized tissues. These adhesion barriers can be divided into categories based on the state of matter and form [10]. A relatively new approach is the use of modified starch-based products, which traditionally have been used solely as hemostats. As hitherto only one starch-based medical device, i.e., 4DryField PH, had been approved for use in adhesion prevention, we made a comparative study with another starch-based hemostat, Arista AH, to find out if starch-based hemostats in general are capable to prevent the formation of adhesions [11]. In that study, we applied both devices in the same manner, i.e., administering the powder and subsequently transforming it into a gel by dripping with saline solution. However, the negative results for Arista AH showed that starch-based hemostats are not generally capable to prevent adhesion formation. As in the meantime two further starch-based hemostats got approved for adhesion formation, we used a challenging and well-reproducible rat model, the recently described Optimized Peritoneal Adhesion Model (OPAM) [12] to test these hemostats, i.e., HaemoCer PLUS and StarSil for their adhesion prevention capability. This model has been shown to induce severest adhesions with high reliability and it has already been used successfully to examine the effectiveness of different anti-adhesive agents in several studies [11, 13, 14]. The model includes abrasion of the cecum and incision of the abdominal wall, as well as meso-stitch approximation of these lesions [12]. Results were compared with published data for 4DryField PH and Arista and saline solution as controls [11].

## Materials and Methods

### Animals

Forty-six male Lewis rats were included in the study. They were housed under standard conditions, had access to fresh water at any time and were fed a standard diet ad libitum. Prior to and after surgery, daily monitoring of body weight and behavioral changes assessed animal welfare. Animal experiments were performed at the central animal laboratory of the Hanover Medical School, Germany, as well as the Therapeutic Experimental Unit, Faculty of Medicine, Nantes, France. All protocols regarding animal life quality were conducted in accordance with national and European regulations. The present study was approved by The Lower Saxony State Office for Consumer Protection and Food Safety (LAVES Hanover, Germany; approval code 12/0751) and the Ethical Committee For Animal Experiments (CEEA) in Pays de la Loire, France (approved under the reference APAFIS 32678).

### Surgical procedures and application of anti-adhesive agents

General anesthesia was achieved by ketamine (80 mg/kg body weight) and xylazine (5 mg/kg body weight) or inhalation of isoflurane 3%. The required level of narcosis was reached when the flexor reflexes were suppressed. A 3 cm long median laparotomy was performed after shaving and sanitizing the abdomen. Adhesion induction was carried out according to the OPAM procedure [12]. First, the cecum was delivered and kept moist with a watery gauze swab, the cecal peritoneum was gently abraded repeatedly over a 1×2 cm area in a standard manner using a dry gauze until removal of visceral peritoneum resulted in sub-serosal bleeding and the creation of a homogenous surface of petechial hemorrhages. Second, the parietal peritoneum and inner muscle layer were sharply dissected to create a 1×2 cm abdominal wall defect. In a third step, both injured areas were approximated using a non-absorbable suture. Prior to surgery, animals were randomly assigned to one of the following four groups: control (n=10), 4DryField PH-treated (n=16), HaemoCer PLUS-treated (n=10) or StarSil-treated (n=10); the latter two groups carried out in Nantes. Control animals received 1.2 ml 0.9% sterile saline solution intraperitoneally. The three anti-adhesive agents 4DryField PH, HaemoCer PLUS and StarSil were each administered in a total amount of 300 mg powder/animal. The powder was evenly distributed on the two defects and then transformed into a gel by dripping with 1.2 ml sterile 0.9% saline solution before the approximating suture was placed. The abdomen was closed using a two-layer closure technique by consecutive sutures. Following surgery, the animals were monitored until they were completely awakened and kept warm using an infrared lamp.

Animals received novaminsulfone and buprenorphine in a body-weight adapted dose to minimize postoperative pain. On postoperative day 7, the animals were sacrificed using CO_2_ narcosis followed by cervical disclosure. The peritoneal cavity was opened by an incision at a left-sided position remote to the original laparotomy scar to prevent damaging any potentially formed adhesions. Specimens of cecum, abdominal wall and adhesions were harvested for histopathological assessment.

### Adhesion assessment

The adhesion formation between the defective abdominal wall and cecum was evaluated macroscopically by two independent observers according to the scoring systems by Lauder et al. [15] and Hoffmann et al. [16]. The Lauder scoring system (Table 1) takes into account number, strength and distribution of adhesions in a single score, while the Hoffmann scoring system (Table 2) consists of three individual scores for area, extent and strength of adhesions that are summed up to yield a total score.

**Table 1.** Adhesion scoring system according to Lauder et al. [15].

| Score | Description |
| --- | --- |
| 0 | No adhesions |
| 1 | Thin filmy adhesions |
| 2 | More than one thin adhesion |
| 3 | Thick adhesion with focal point |
| 4 | Thick adhesion with planar attachment |
| 5 | Very thick vascularized adhesions or more than one planar adhesion |

**Table 2.** Adhesion scoring system according to Hoffmann et al. [16].

| Score | Description |
| --- | --- |
| Area score |  |
| 0 | No adhesion |
| 1 | Cecum to bowel adhesion |
| 2 | Cecum to sidewall adhesion over less than 25% of the abraded surface area |
| 3 | Cecum to sidewall adhesion between 25 and 50% of the abraded surface area |
| 4 | Cecum to sidewall adhesion over more than 50% of the surface area |
| Strength score |  |
| 0 | No adhesion |
| 1 | Gentle traction required to break adhesion |
| 2 | Blunt dissection required to break adhesion |
| 3 | Sharp dissection required to break adhesion |
| Extent score |  |
| 0 | No adhesion |
| 1 | Filmy adhesion |
| 2 | Vascularized adhesion |
| 3 | Opaque or cohesive adhesion |

### Statistical analyses

Adhesion scores are presented as arithmetic means with standard deviations (SD). Since some of the data sets did not follow a Gaussian distribution (as determined using the D’Agostino-Pearson normality test), the multiple comparisons of adhesion scores of the four groups were performed using Kruskal-Wallis test followed by Dunn’s multiple comparisons test for non-parametric data (which utilizes correction for multiple comparison by statistical hypothesis testing). Groups were defined to be significantly different if p<0.05. Statistical analyses were performed using GraphPad Prism (Version 7.0b for Mac OS, GraphPad Software, Inc., La Jolly, USA).

### Histology

Surgical specimens were fixed in buffered 4% formaldehyde solution. After dehydration and paraffin embedding, serial thin sections of 1–2 μm were mounted on glass slides, stained with standard Hematoxylin and Eosin (HE), Elastika-van-Gieson (EvG) and periodic acid-Schiff (PAS) staining (Sigma Aldrich Co Ltd, USA) and light microscope examinations were performed by experienced pathologists.

The quantitative analysis of the histologic stainings was performed using Zühlke’s microscopic adhesion classification (Table 3).

**Table 3.** Microscopic adhesion classification according to Zühlke et al. [17].

| Score | Description |
| --- | --- |
| 0 | No adhesions |
| 1 | Weak connective tissue, rich cell, new and old fibrin, thin reticulin fibrils |
| 2 | Connective tissue which has cells and capillaries. few collagen fibers |
| 3 | Thicker connective tissue. Few cells and elastic and smooth muscle fibers, more vessels |
| 4 | Old and thick granulation tissue, poor cells, difficult separation of serosal surfaces |

This system has already been established for grading of peritoneal adhesions induced with models very similar to OPAM [18, 19], as well as in a recent OPAM study comparing the efficacy of 4DryField PH vs. Arista AH [11].

## Results

All animals showed a comparable course of viability and body weight. None of the animals had to be sacrificed prematurely due to complications, all 46 animals completed the study.

### Adhesion development

As published earlier, in the control group, 9 of 10 animals showed peritoneal adhesions, which were rated with the maximum Lauder score, as well as the maximum scores regarding all Hoffmann categories (Figure 1A). None of the sixteen 4DryField PH-treated animals developed any adhesions (Figure 1B) [11]. In contrast, all HaemoCer PLUS-treated (Figure 1C) and all StarSil-treated (Figure 1D) animals developed peritoneal adhesions. In these two groups, Lauder score ranged from 0.5 to 4, respectively. Hoffmann Area scores ranged from 0.5 to 3.5 in the HaemoCer PLUS group and from 0.5 to 3 in the StarSil group. Hoffmann Strength scores ranged from 0.5 to 3.5 in the HaemoCer PLUS group and from 0.5 to 2.5 in the StarSil group. Hoffmann Extent scores ranged from 0.5 to 3.5 in the HaemoCer PLUS group and from 0.5 to 3 in the StarSil group. The mean score value of each group was calculated and tested for significant differences (Table 4). Herein, 4DryField PH reduced the incidence and severity of peritoneal adhesion formation significantly compared to the control, as well as to the HaemoCer PLUS- and StarSil-treated groups. In contrast, HaemoCer PLUS- and StarSil-treatment did not lead to a statistically significant reduction of adhesion formation in comparison to control animals and when compared with each other.

**Table 4.** Arithmetic mean values (AM), standard deviations (SD) and p-values in comparison to the control (p (ctrl)), 4DryField PH (p (4DF)) or HaemoCer PLUS (p (HCP)) groups (statistically significant difference if p<0.05, ^*^).

| Score | Group | AM | SD | p (ctrl) | p (4DF) | p (HCP) |
| --- | --- | --- | --- | --- | --- | --- |
| Lauder | Control | 4.5 | 1.6 |  |  |  |
|  | 4DryField PH | 0.0 | 0.0 | <0.0001* |  |  |
|  | HaemoCer PLUS | 1.3 | 1.1 | 0.163 | 0.0079* |  |
|  | StarSil | 2.2 | 1.4 | 0.6039 | 0.0007* | >0.9999 |
| Hoffmann Area | Control | 3.6 | 1.3 |  |  |  |
|  | 4DryField PH | 0.0 | 0.0 | <0.0001* |  |  |
|  | HaemoCer PLUS | 1.1 | 0.9 | 0.1323 | 0.0104* |  |
|  | StarSil | 1.7 | 0.9 | 0.7034 | 0.0005* | >0.9999 |
| Hoffmann Strength | Control | 2.7 | 0.9 |  |  |  |
|  | 4DryField PH | 0.0 | 0.0 | <0.0001* |  |  |
|  | HaemoCer PLUS | 1.0 | 0.9 | 0.1653 | 0.0167* |  |
|  | StarSil | 1.6 | 0.7 | >0.9999 | 0.0005* | >0.9999 |
| Hoffmann Extent | Control | 2.7 | 0.9 |  |  |  |
|  | 4DryField PH | 0.0 | 0.0 | <0.0001* |  |  |
|  | HaemoCer PLUS | 1.1 | 0.9 | 0.3094 | 0.0067* |  |
|  | StarSil | 1.7 | 0.9 | >0.9999 | 0.0003* | >0.9999 |
| Hoffmann Total | Control | 9.0 | 3.2 |  |  |  |
|  | 4DryField PH | 0.0 | 0.0 | <0.0001* |  |  |
|  | HaemoCer PLUS | 3.1 | 2.7 | 0.2497 | 0.0074* |  |
|  | StarSil | 5.0 | 2.5 | >0.9999 | 0.0004* | >0.9999 |

**Figure 1.**
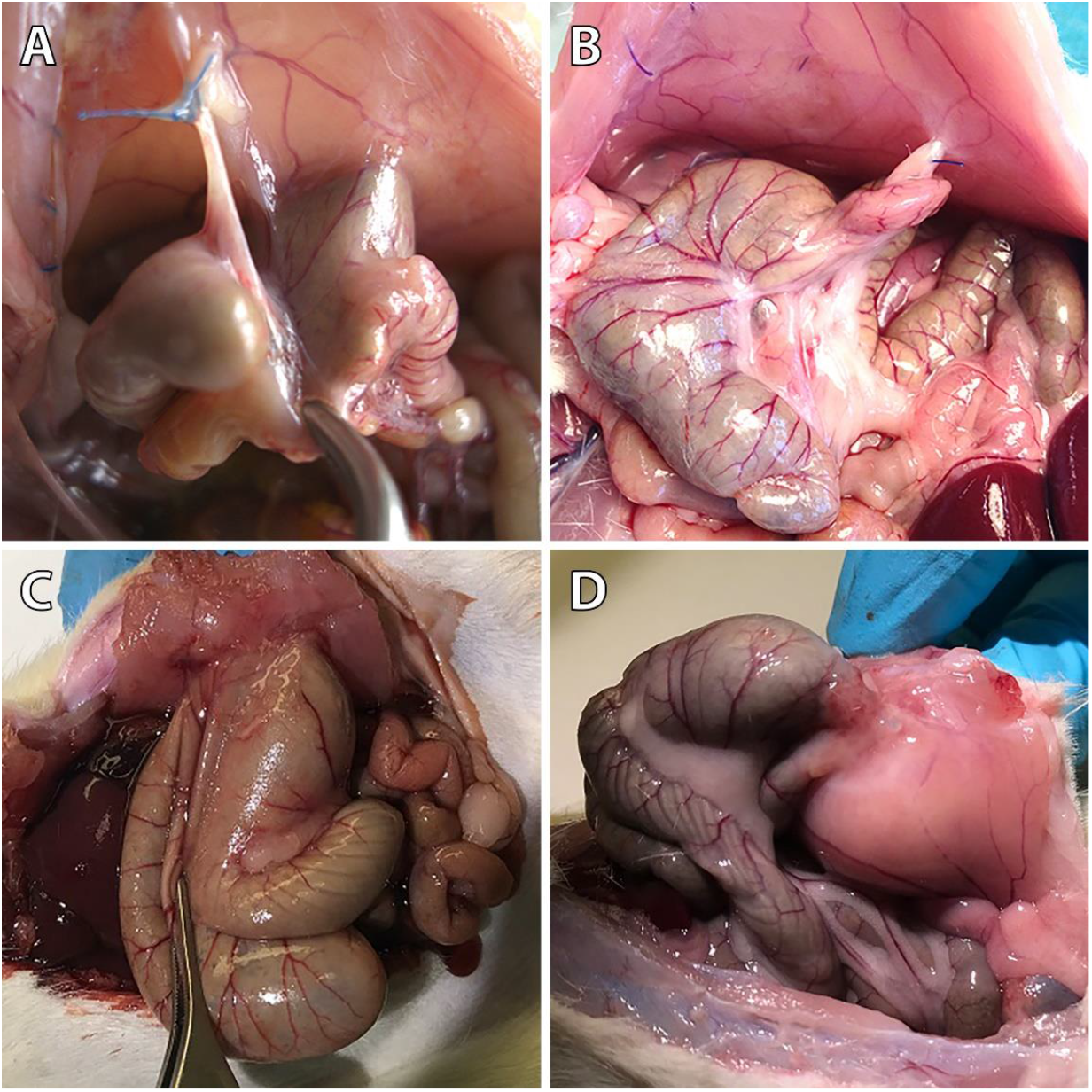
Representative photographs of the pathological evaluation of control (A), 4DryField PH-(B), HaemoCer PLUS-(C), and StarSil-(D) treated rats on day 7.

### Histology

Figure 2 shows representative PAS-stained tissue slides from all four groups. Figure 2A shows a control animal where the smooth muscle layers of the cecum (top) are fused to skeletal muscles of the abdominal wall (bottom) via dense granulating tissue. The histological findings support the macroscopic observation that both, cecum and abdominal wall, could not readily be separated by mechanical force. Figure 2B shows abdominal wall and Figure 2C cecal tissue of an animal from the 4DryField PH group. In contrast to 9 of the 10 control animals, no agglutinations occurred in the 4DryField PH group. Furthermore, in all animals of the 4DryField PH group the lesions of the cecum and the abdominal wall defect already healed, and both featured neomesothelial cell coverage. The former abdominal wall defect was filled with fibrous tissue, which still contained slight remnants of 4DryField PH particles. These data had already been published [11]. Figure 2D shows a representative photograph of a HaemoCer PLUS-treated animal. As in Figure 2A, the smooth muscles of the cecum (top) were fused to the skeletal muscles of the abdominal wall (bottom) via dense granulation tissue, preventing separation of cecum and abdominal wall by mechanical force. In contrast to 4DryField PH, remnants of HaemoCer PLUS were not found in any of the histological slides. Similarly, Figure 2E shows PAS staining of a representative animal from the StarSil group, in which the smooth muscles of the cecum (top) were fused to the skeletal muscles of the abdominal wall (bottom) via dense granulation tissue, preventing separation of cecum and abdominal wall by mechanical force. As in HaemoCer PLUS and in contrast to 4DryField PH, remnants of StarSil were not found in any of the histological slides.

**Figure 2.**
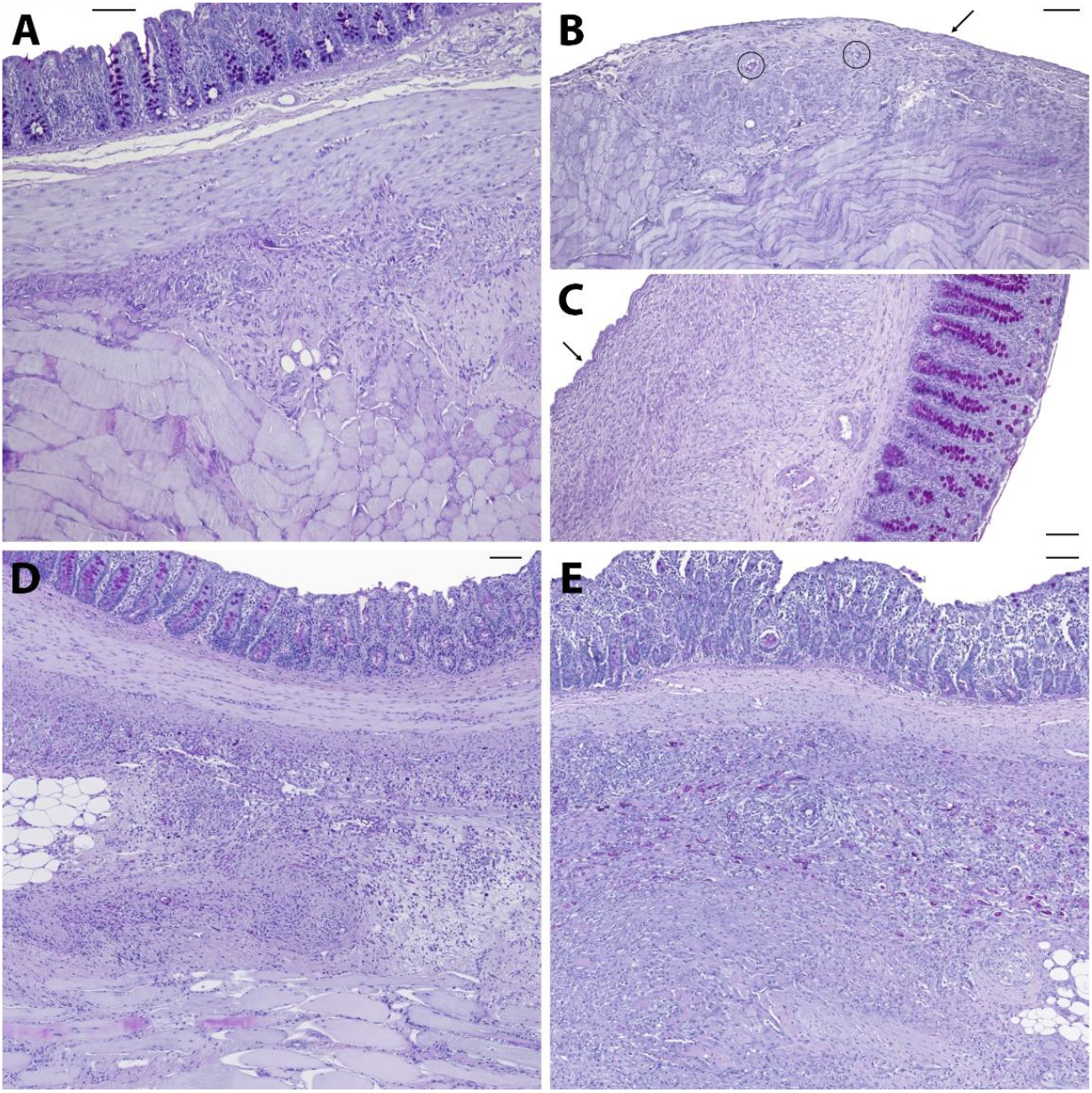
Representative histological slides (PAS-stained) of animals from the control (A), 4DryField PH-(B, C), HaemoCer PLUS-(D), and StarSil-(E) treated groups. Black arrows indicate neomesothelial coverage, black circles denote remnants of 4DryField PH. Scale bars: 100 μm.

The microscopic classification of the adhesions according to Zühlke et al. [17] was performed for all animals. In the control group, one animal was scored 0, two were scored 3 and seven were scored 4. In the 4DryField PH group, the microscopic assessment was equivalent to the macroscopic investigation with all 16 animals being scored 0. In the HaemoCer PLUS group, three animals were scored 0, two were scored 2, three were scored 3, and two were scored 4. In the StarSil group, three animals were scored 0, one was scored 2, one was scored 3, and five were scored 4.

Like for the macroscopic adhesion assessment, the mean scores were calculated and tested for significant differences (Table 5). The results conform with the macroscopic assessment and 4DryField PH-treated animals achieved significantly lower mean Zühlke scores than HaemoCer PLUS- and StarSil-treated ones as well as controls. In contrast, HaemoCer PLUS- and StarSil-treatment did not lead to a statistically significant reduction of Zühlke adhesion scores in comparison to control animals and when compared among themselves.

**Table 5.** Arithmetic mean values (AM), standard deviations (SD) and p-values in comparison to the control (p (ctrl)), 4DryField PH (p (4DF)) or HaemoCer PLUS (p (HCP)) groups (statistically significant difference if p<0.05, ^*^).

| Score | Group | AM | SD | p (ctrl) | p (4DF) | p (HCP) |
| --- | --- | --- | --- | --- | --- | --- |
| Zühlke | Control | 3.3 | 1.3 |  |  |  |
|  | 4DryField PH | 0.0 | 0.0 | **<0.0001\*** |  |  |
|  | HaemoCer PLUS | 2.1 | 1.6 | 0.5305 | **0.0436\*** |  |
|  | StarSil | 2.5 | 1.8 | >0.9999 | **0.0044\*** | >0.9999 |

## Discussion

As shown in previous studies [11-14], the OPAM consistently induced severe peritoneal adhesions after cecal abrasion and creation of abdominal wall defects in rats.

4DryField PH revealed excellent adhesion prevention capabilities, preventing the formation of any adhesions. In contrast, HaemoCer PLUS and StarSil did not lead to a statistically significant reduction of adhesion formation in comparison to control animals and when compared among themselves. Additionally, the adhesion prevention efficacy of HaemoCer PLUS and StarSil was significantly lower than in 4DryField PH. Although surgeries were performed at two study centers, limited comparability of the results arising from differing surgical performance can be ruled out due to strict monitoring of the comparability as described above. Additionally, the OPAM has been used at the Hanover Medical School extensively (12-14) and different surgeons have performed surgeries following this protocol in the past, but a correlation of results with the respective surgeon has never been observed. Correspondingly, the model has been shown to be highly reliable and very robust.

In previous studies, 4DryField PH was highly effective in preventing peritoneal adhesions when it had been applied prophylactically either as a premixed gel or as powder that was transformed *in situ* into a gel by adding saline solution [13, 14]. For HaemoCer PLUS and StarSil, this is the first experimental study examining their adhesion prevention capabilities. For HaemoCer PLUS, the efficacy in reducing the formation of adhesions was previously investigated in a clinical trial on 107 patients undergoing open and laparoscopic gynecological surgeries [10, 20]. In this study, HaemoCer PLUS was found to be ineffective in preventing adhesions as confirmed in second look operations [20]. The application mode of HaemoCer PLUS in this study was an *in-situ* gel [20]. For StarSil, no previous studies examining its adhesion prevention capabilities have been found.

Considering the available data on the adhesion prevention capabilities of various starch-based powder hemostats, only 4DryField PH has shown effective adhesion prevention, whereas no efficacy has been detected for HaemoCer PLUS and StarSil (both certified for adhesion prevention), as well as Arista AH (not certified for adhesion prevention) (11). This has been evidenced macroscopically by using both Lauder and Hoffmann scoring systems, as well as microscopically by systematic histopathological examinations using the Zühlke microscopic classification system in this rat model. The microscopic analyses showed no agglutinations of cecum and abdominal wall in the 4DryField PH group, whereas most animals had tight agglutinations of cecum and abdominal wall via granulating tissue in the HaemoCer PLUS, StarSil and Arista AH groups, comparable to those of the control animals.

Based on the differences in efficacy observed between 4DryField PH and Arista AH, we had concluded that starch-based powder hemostats do not generally have the capability to function as effective adhesion prevention devices [11]. Considering the negative results obtained for two further starch-based hemostats in the present study, i.e., HaemoCer PLUS and StarSil, this assumption is confirmed. Krämer et al. [10] assumed that the short retention time of only about 1–3 days of starch-based hemostats such as Arista and HaemoCer PLUS constitutes an important reason for their reduced efficacy. The results of the present study further support this hypothesis as the retention times of Arista AH, HaemoCer PLUS and StarSil are distinctly shorter than that of 4DryField PH (1 –3 days vs. 7 days). In addition, these products differ in other characteristics, such as water absorption. All these data show that various starch-based hemostats indeed exhibit significant differences in product properties. Accordingly, starch-based powder hemostats are not naturally capable to reduce adhesion formation. Instead, the effectiveness depends on the specific properties of the individual product, which are commonly not reported in detail and might be of interest for further investigations.

## Conclusions

Only 4DryField PH but neither HaemoCer PLUS nor StarSil were capable to effectively prevent adhesion formation, corroborating the assumption that starch-based hemostats do not generally have the capability to act as effective adhesion prevention devices.

## Data Availability

All data produced in the present work are contained in the manuscript

## Acknowledgments

The authors would like to thank Dres. Valérie Dumay and Pierre Layrolle for generous sharing of data obtained in the Therapeutic Experimental Unit, Faculty of Medicine, Nantes, France. They performed the HaemoCer PLUS and StarSil experiments, funded by PlantTec Medical GmbH, Germany. The authors are also grateful to Valentina Osmani for editing the manuscript.

## Competing Interests

The authors have declared that no competing interest exists.

## Notes

### Competing Interest Statement

The authors have declared no competing interest.

### Funding Statement

This study did not receive any funding

